# Are metastatic tumours histologically similar to the primary? Result of a blinded experiment

**DOI:** 10.1101/2023.02.06.23285511

**Authors:** Akshita Singh, Nirmala Jambhekar, Tanuja Shet, Mukta Ramadwar, Rajesh Dikshit, Indraneel Mittra

**Author notes:** Author for correspondence: Indraneel Mittra.

## Abstract

There is an entrenched belief among pathologists, biologists and clinicians alike that metastatic tumours are histologically similar to the primary. Although this belief has never been scientifically tested, it forms the conceptual foundation upon which metastasis research has been based for decades. In order to scientifically test this dogma, we conducted a blinded experiment in which three highly experienced professor grade cancer pathologists were asked to predict the site of origin of 298 metastatic tumours arising at nine different sites. The accuracy of prediction of all three examiners was just under 50%. However, there was very little inter-observer agreement, and Kappa statistic generated the value of 0.27 which is much below the figure of 0.4 required to detect even a moderate level of concordance. In only 56 / 298 (18.8%) cases did the examiners agree with each other’s diagnosis. Thus the possibility cannot be excluded that the correct diagnoses made by the examiners had happened by play of chance. Our finding suggests that histological relationship between primary and metastatic tumours requires further investigation.

## Introduction

There is a widely held belief among pathologists, biologists and clinicians alike that metastatic tumours are histologically similar to the primary. Although this theory has never been scientifically tested, it has formed the conceptual foundation upon which research on cancer metastasis has been based for decades. This theory has led to the formulation of the second theory which posits that cells which exit the primary tumour must themselves travel to distant organs and colonize to form secondary tumours such that they resemble the primary. Yet, after decades of research the mechanistic basis of the metastatic cascade has not been satisfactorily elucidated^1,2^. An alternative line of research has suggested that nucleic acids derived from dying cancer cells that are carried via blood stream to distant organs are the instigators of metastasis by their ability to transform cells of the target organs to generate new tumours that masquerade as metastasis^3–8^. If this theory were to be true, secondary tumours should not be histologically similar to the primary. We undertook a study to blindly test the veracity of the two theories of metastasis.

## Materials and Methods

We retrieved 298 paraffin blocks of metastatic tumours from the archives of the Pathology Department of Tata Memorial Hospital. The blocks comprised of metastatic tumours arising at 9 different sites viz. brain, bone, lung, liver, cutaneous nodules, inguinal lymph node, mediastinal lymph node, mesenteric lymph node and supra-clavicular lymph node. These metastatic tumours had arisen from primary tumours at 22 different sites (Table 1). The paraffin blocks were sectioned and fresh histological slides were prepared and stained with H&E. The slides were blindly coded. Three highly experienced professor grade cancer pathologists with at least 20 years of professional experience were asked to identify the primary sites of origin of the metastatic tumours. The three examiners are co-correspondents of this letter. Of the 298 slides, 110 were examined by all three examiners while 188 slides were examined by two examiners. The predictions made by the examiners were tabulated against the appropriate primary sites and the accuracy of prediction was recorded (Table 2). Inter-examiner concordance was determined by Kappa statistics.

**Table 1:** Metastatic tumours and their primary sites of origin

| Metastatic sites<br>→ | Bone | Brain | Lung | Liver | Cutaneous | Inguinal LN | Mediastinal<br>LN | Mesenteric<br>LN | Supraclavicular LN |
| --- | --- | --- | --- | --- | --- | --- | --- | --- | --- |
| Primary sites | Thyroid (n=7) | Breast (n=12) | Breast (n=19) | Breast (n=21) | Breast (n=24) | Ovary (n=10) | Lung (n=24) | Colorectal (n=19) | Breast (n=21) |
|  | Breast (n=6) | Lung (n=9) | Renal (n=11) | Colorectal (n=8) | Lung (n=2) | Colorectal (n=5) | Esophagus (n=7) | Endometrium (n=8) | Lung (n=3) |
|  | Lung (n=1) | Thyroid (n=2) | Colorectal (n=8) | Ovary (n=6) | Colon (n=2) | Renal (n=2) | Thyroid (n=5) | Ovary (n=7) | Ovary (n=3) |
|  | Renal (n=1) | Ovary (n=2) | Parotid (n=2) | Periampullary/pancreas (n=5) | Ovary (n=1) | Breast (n=2) | Breast (n=1) | Jejunum (n=2) | Thyroid (n=1) |
|  | Prostate (n=1) | Sebaceous gland (n=1) | Urinary bladder (n=1) | Biliary tract (n=3) | Salivary gland (n=1) | Prostate (n=2) |  | Periampullary/pancreas (n=1) | Stomach (n=1) |
|  | Colorectal (n=1) |  | Thyroid (n=1) | Renal (n=2) | Thyroid (n=1) | Vulva (n=1) |  |  | Renal (n=1) |
|  |  |  |  | Lung (n=2) |  | Vagina (n=1) |  |  | Periampullary/pancreas (n=1) |
|  |  |  |  | Stomach (n=2) |  | Ileum (n=1) |  |  |  |
|  |  |  |  | Urinary Bladder (n=1) |  |  |  |  |  |
|  |  |  |  | Duodenum (n=1) |  |  |  |  |  |
|  |  |  |  | Esophagus (n=1) |  |  |  |  |  |
|  |  |  |  | Thyroid (n=1) |  |  |  |  |  |

**Table 2.** Accuracy of diagnosis.

| Metastatic sites | Sr. No. | 1st examiner | 2nd examiner | 3rd examiner | Score |
| --- | --- | --- | --- | --- | --- |
| BONE | 1 | 0 | 0 | 1 | 1/3 |
|  | 2 | 1 | 0 | 1 | 2/3 |
|  | 3 | 1 | 0 | NE | 1/2 |
|  | 4 | 1 | 1 | 0 | 2/3 |
|  | 5 | 1 | 1 | 1 | 3/3 |
|  | 6 | 1 | 1 | 1 | 3/3 |
|  | 7 | 1 | 1 | 1 | 3/3 |
|  | 8 | 1 | 1 | 1 | 3/3 |
|  | 9 | 0 | 0 | 0 | 0/3 |
|  | 10 | 0 | 0 | 1 | 1/3 |
|  | 11 | 1 | 0 | 1 | 2/3 |
|  | 12 | 1 | 0 | NE | 1/2 |
|  | 13 | 1 | 0 | 1 | 2/3 |
|  | 14 | 1 | 1 | 1 | 3/3 |
|  | 15 | 1 | 1 | 1 | 3/3 |
|  | 16 | NE | 1 | 1 | 2/2 |
|  | 17 | 1 | 0 | 0 | 1/3 |
| BRAIN | 18 | 1 | 1 | 1 | 3/3 |
|  | 19 | 1 | 0 | NE | 1/2 |
|  | 20 | 0 | 0 | NE | 0/2 |
|  | 21 | 1 | 1 | 0 | 2/3 |
|  | 22 | 0 | 1 | 1 | 2/3 |
|  | 23 | 0 | 0 | 0 | 0/3 |
|  | 24 | 0 | 0 | 1 | 1/3 |
|  | 25 | 1 | 0 | 1 | 2/3 |
|  | 26 | 0 | 1 | 0 | 1/3 |
|  | 27 | 0 | 0 | NE | 0/2 |
|  | 28 | 0 | 1 | 0 | 1/3 |
|  | 29 | 0 | 1 | 1 | 2/3 |
|  | 30 | 0 | 0 | 0 | 0/3 |
|  | 31 | 1 | 1 | 0 | 2/3 |
|  | 32 | 0 | 0 | NE | 0/2 |
|  | 33 | 0 | 0 | 0 | 0/3 |
|  | 34 | 0 | 1 | 1 | 2/3 |
|  | 35 | 1 | 1 | 1 | 3/3 |
|  | 36 | 1 | 0 | 0 | 1/3 |
|  | 37 | 1 | 1 | 1 | 3/3 |
|  | 38 | 1 | 1 | 0 | 2/3 |
|  | 39 | 0 | 0 | 1 | 1/3 |
|  | 40 | 0 | 1 | 0 | 1/3 |
|  | 41 | 0 | 0 | 0 | 0/3 |
|  | 42 | 0 | 0 | 0 | 0/3 |
|  | 43 | 0 | 1 | 1 | 2/3 |

**1 = Correct diagnosis; 0 = Incorrect diagnosis; NE = Not Examined; Yellow lines = Perfect agreement.**
| Metastatic sites | Sr. No. | 1st examiner | 2nd examiner | 3rd examiner | Score |
| --- | --- | --- | --- | --- | --- |
| CUTANEOUS | 44 | NE | 1 | 1 | 2/2 |
|  | 45 | NE | 1 | 0 | 1/2 |
|  | 46 | NE | NE | 1 | 1/1 |
|  | 47 | NE | 0 | 0 | 0/2 |
|  | 48 | NE | 1 | 0 | 1/2 |
|  | 49 | NE | 0 | 1 | 1/2 |
|  | 50 | NE | 0 | 1 | 1/2 |
|  | 51 | NE | NE | 1 | 1/1 |
|  | 52 | NE | 1 | 0 | 1/2 |
|  | 53 | NE | 0 | 0 | 0/2 |
|  | 54 | NE | 0 | 1 | 1/2 |
|  | 55 | NE | 1 | 1 | 2/2 |
|  | 56 | NE | 1 | 0 | 1/2 |
|  | 57 | NE | 1 | 0 | 1/2 |
|  | 58 | NE | 1 | NE | 1/1 |
|  | 59 | NE | NE | 1 | 1/1 |
|  | 60 | NE | 1 | 0 | 1/2 |
|  | 61 | NE | 0 | NE | 0/1 |
|  | 62 | NE | 0 | 0 | 0/2 |
|  | 63 | NE | NE | 0 | 0/1 |
|  | 64 | NE | 0 | 0 | 0/2 |
|  | 65 | NE | 0 | 0 | 0/2 |
|  | 66 | NE | 0 | 0 | 0/2 |
|  | 67 | NE | 0 | 1 | 1/2 |
|  | 68 | NE | 1 | 0 | 1/2 |
|  | 69 | NE | 0 | 0 | 0/2 |
|  | 70 | NE | NE | 1 | 1/1 |
|  | 71 | NE | 0 | 1 | 1/2 |
|  | 72 | NE | 1 | 0 | 1/2 |
|  | 73 | NE | 0 | 1 | 1/2 |
| INGUINAL LN | 74 | 0 | NE | 0 | 0/2 |
|  | 75 | 0 | 0 | 0 | 0/3 |
|  | 76 | 1 | 1 | 1 | 3/3 |
|  | 77 | 0 | 0 | 0 | 0/3 |
|  | 78 | 1 | 0 | 0 | 1/3 |
|  | 79 | 0 | 0 | 0 | 0/3 |
|  | 80 | 0 | NE | 0 | 0/2 |
|  | 81 | 1 | NE | 0 | 1/2 |
|  | 82 | 1 | 0 | 0 | 1/3 |
|  | 83 | 0 | 0 | 0 | 0/3 |
|  | 84 | 0 | NE | 1 | 1/2 |
|  | 85 | 0 | 1 | 1 | 2/3 |
|  | 86 | 0 | 0 | 0 | 0/3 |
|  | 87 | 0 | 1 | 1 | 2/3 |
|  | 88 | 0 | 0 | 0 | 0/3 |
|  | 89 | 0 | 1 | 0 | 1/3 |
|  | 90 | 1 | 1 | 1 | 3/3 |
|  | 91 | 1 | 1 | 1 | 3/3 |
|  | 92 | 0 | 0 | 0 | 0/3 |

**1 = Correct diagnosis; 0 = Incorrect diagnosis; NE = Not Examined; Yellow lines = Perfect agreement.**
| Metastatic sites | Sr. No. | 1st examiner | 2nd examiner | 3rd examiner | Score |
| --- | --- | --- | --- | --- | --- |
|  | 93 | 1 | 1 | 1 | 3/3 |
|  | 94 | 0 | 1 | 0 | 1/3 |
|  | 95 | 0 | NE | 0 | 0/2 |
|  | 96 | 1 | 1 | 1 | 3/3 |
|  | 97 | 0 | 0 | 1 | 1/3 |
| LIVER | 98 | 1 | 1 | 0 | 2/3 |
|  | 99 | 0 | 1 | 1 | 2/3 |
|  | 100 | 0 | 0 | 0 | 0/3 |
|  | 101 | 0 | 1 | 0 | 1/3 |
|  | 102 | 0 | 1 | 0 | 1/3 |
|  | 103 | 1 | 1 | 0 | 2/3 |
|  | 104 | 0 | 0 | 0 | 0/3 |
|  | 105 | 0 | 1 | 0 | 1/3 |
|  | 106 | 0 | 1 | 0 | 1/3 |
|  | 107 | 0 | 1 | 0 | 1/3 |
|  | 108 | NE | NE | 1 | 1/1 |
|  | 109 | 0 | 0 | 0 | 0/3 |
|  | 110 | 0 | 0 | 0 | 0/3 |
|  | 111 | 0 | 0 | 0 | 0/3 |
|  | 112 | 1 | 1 | 1 | 3/3 |
|  | 113 | 0 | 0 | 0 | 0/3 |
|  | 114 | 0 | 0 | 0 | 0/3 |
|  | 115 | 0 | 1 | 0 | 1/3 |
|  | 116 | NE | 1 | 1 | 2/2 |
|  | 117 | NE | 0 | 0 | 0/2 |
|  | 118 | NE | 0 | 0 | 0/2 |
|  | 119 | NE | 0 | 1 | 1/2 |
|  | 120 | NE | 1 | 0 | 1/2 |
|  | 121 | NE | 0 | 0 | 0/2 |
|  | 122 | NE | 1 | 1 | 2/2 |
|  | 123 | NE | 0 | 0 | 0/2 |
|  | 124 | NE | 0 | 0 | 0/2 |
|  | 125 | NE | 1 | 1 | 2/2 |
|  | 126 | NE | 0 | 0 | 0/2 |
|  | 127 | NE | 0 | 1 | 1/2 |
|  | 128 | NE | 1 | 0 | 1/2 |
|  | 129 | NE | 0 | 0 | 0/2 |
|  | 130 | NE | 1 | 0 | 1/2 |
|  | 131 | NE | 0 | 0 | 0/2 |
|  | 132 | NE | 1 | 1 | 2/2 |
|  | 133 | NE | 0 | 0 | 0/2 |
|  | 134 | NE | 0 | 0 | 0/2 |
|  | 135 | NE | 0 | NE | 0/1 |
|  | 136 | NE | 0 | 0 | 0/2 |
|  | 137 | NE | NE | 1 | 1/1 |
|  | 138 | NE | 0 | 0 | 0/2 |
|  | 139 | NE | 0 | 0 | 0/2 |
|  | 140 | NE | 1 | 1 | 2/2 |
|  | 141 | NE | 1 | 1 | 2/2 |

**1 = Correct diagnosis; 0 = Incorrect diagnosis; NE = Not Examined; Yellow lines = Perfect agreement.**
| Metastatic sites | Sr. No. | 1st examiner | 2nd examiner | 3rd examiner | Score |
| --- | --- | --- | --- | --- | --- |
|  | 142 | NE | NE | 0 | 0/1 |
|  | 143 | NE | 0 | 0 | 0/2 |
|  | 144 | NE | 1 | 0 | 1/2 |
|  | 145 | NE | NE | 0 | 0/1 |
|  | 146 | NE | 0 | 0 | 0/2 |
|  | 147 | NE | 0 | 1 | 1/2 |
|  | 148 | NE | 1 | 0 | 1/2 |
|  | 149 | NE | 1 | 1 | 2/2 |
|  | 150 | NE | NE | 0 | 0/1 |
| LUNG | 151 | 1 | 1 | 1 | 3/3 |
|  | 152 | 0 | NE | 1 | 1/2 |
|  | 153 | 0 | 1 | 0 | 1/3 |
|  | 154 | 0 | NE | NE | 0/1 |
|  | 155 | 0 | 0 | 1 | 1/3 |
|  | 156 | 1 | NE | 1 | 2/2 |
|  | 157 | 0 | 1 | 1 | 2/3 |
|  | 158 | 0 | 0 | 0 | 0/3 |
|  | 159 | 1 | 1 | 1 | 3/3 |
|  | 160 | 0 | 0 | 1 | 1/3 |
|  | 161 | 0 | 1 | 0 | 1/3 |
|  | 162 | 1 | 1 | 0 | 2/3 |
|  | 163 | 1 | 1 | 0 | 2/3 |
|  | 164 | 0 | 0 | 0 | 0/3 |
|  | 165 | 0 | 1 | 0 | 1/3 |
|  | 166 | 0 | NE | 1 | 1/2 |
|  | 167 | 0 | 0 | 0 | 0/3 |
|  | 168 | 1 | 1 | 0 | 2/3 |
|  | 169 | 0 | 0 | 0 | 0/3 |
|  | 170 | 0 | 0 | 0 | 0/3 |
|  | 171 | 1 | 1 | 1 | 3/3 |
|  | 172 | 0 | 1 | 1 | 2/3 |
|  | 173 | 0 | 0 | 0 | 0/3 |
|  | 174 | 0 | 0 | 0 | 0/3 |
|  | 175 | 0 | 0 | 0 | 0/3 |
|  | 176 | 0 | 1 | 1 | 2/3 |
|  | 177 | 0 | 1 | 0 | 1/3 |
|  | 178 | 1 | 0 | 1 | 2/3 |
|  | 179 | 0 | 1 | 1 | 2/3 |
|  | 180 | 1 | 1 | 1 | 3/3 |
|  | 181 | 1 | 0 | 1 | 2/3 |
|  | 182 | 0 | 0 | 1 | 1/3 |
|  | 183 | 0 | 0 | 0 | 0/3 |
|  | 184 | 1 | 0 | 1 | 2/3 |
|  | 185 | 0 | 0 | 0 | 0/3 |
|  | 186 | 1 | 1 | 1 | 3/3 |
|  | 187 | 1 | 1 | 1 | 3/3 |
|  | 188 | 0 | 1 | 1 | 2/3 |
|  | 189 | 1 | 0 | 0 | 1/3 |
|  | 190 | 0 | 0 | 0 | 0/3 |

**1 = Correct diagnosis; 0 = Incorrect diagnosis; NE = Not Examined; Yellow lines = Perfect agreement.**
| Metastatic sites | Sr. No. | 1st examiner | 2nd examiner | 3rd examiner | Score |
| --- | --- | --- | --- | --- | --- |
|  | 191 | 1 | 1 | 0 | 2/3 |
|  | 192 | 0 | 0 | 1 | 1/3 |
| MEDIASTINAL LN | 193 | NE | NE | 1 | 1/1 |
|  | 194 | NE | 1 | 1 | 2/2 |
|  | 195 | NE | 1 | 1 | 2/2 |
|  | 196 | NE | 0 | 0 | 0/2 |
|  | 197 | NE | 0 | NE | 0/1 |
|  | 198 | NE | 1 | 1 | 2/2 |
|  | 199 | NE | 1 | 0 | 1/2 |
|  | 200 | NE | 0 | 1 | 1/2 |
|  | 201 | NE | 0 | 0 | 0/2 |
|  | 202 | NE | NE | 1 | 1/1 |
|  | 203 | NE | 0 | 0 | 0/2 |
|  | 204 | NE | 0 | 0 | 0/2 |
|  | 205 | NE | 0 | 0 | 0/2 |
|  | 206 | NE | 0 | 1 | 1/2 |
|  | 207 | NE | 0 | NE | 0/1 |
|  | 208 | NE | 0 | 0 | 0/2 |
|  | 209 | NE | NE | 1 | 1/1 |
|  | 210 | NE | 0 | 0 | 0/2 |
|  | 211 | NE | 0 | 1 | 1/2 |
|  | 212 | NE | 0 | 0 | 0/2 |
|  | 213 | NE | 0 | 1 | 1/2 |
|  | 214 | NE | 0 | 0 | 0/2 |
|  | 215 | NE | 1 | 0 | 1/2 |
|  | 216 | NE | 1 | 1 | 2/2 |
|  | 217 | NE | 0 | 1 | 1/2 |
|  | 218 | NE | 0 | 0 | 0/2 |
|  | 219 | NE | 1 | 1 | 2/2 |
|  | 220 | NE | 0 | 1 | 1/2 |
|  | 221 | NE | 0 | 0 | 0/2 |
|  | 222 | NE | 0 | 0 | 0/2 |
|  | 223 | NE | 1 | 1 | 2/2 |
|  | 224 | NE | 0 | 1 | 1/2 |
|  | 225 | NE | 0 | 1 | 1/2 |
|  | 226 | NE | 1 | 1 | 2/2 |
|  | 227 | NE | 1 | 1 | 2/2 |
|  | 228 | NE | 0 | 0 | 0/2 |
|  | 229 | NE | 0 | 1 | 1/2 |
| MESENEERIC LN | 230 | NE | 0 | 0 | 0/2 |
|  | 231 | NE | 1 | 0 | 1/2 |
|  | 232 | NE | 1 | 0 | 1/2 |
|  | 233 | NE | 1 | 1 | 2/2 |
|  | 234 | NE | 0 | 0 | 0/2 |
|  | 235 | NE | 1 | 1 | 1/2 |
|  | 236 | NE | 0 | 0 | 0/2 |
|  | 237 | NE | 0 | 0 | 0/2 |
|  | 238 | NE | 1 | 1 | 2/2 |
|  | 239 | NE | 1 | 1 | 2/2 |

**1 = Correct diagnosis; 0 = Incorrect diagnosis; NE = Not Examined; Yellow lines = Perfect agreement.**
| Metastatic sites | Sr. No. | 1st examiner | 2nd examiner | 3rd examiner | Score |
| --- | --- | --- | --- | --- | --- |
|  | 240 | NE | 1 | 0 | 1/2 |
|  | 241 | NE | 1 | 0 | 1/2 |
|  | 242 | NE | 1 | 1 | 2/2 |
|  | 243 | NE | 0 | 0 | 0/2 |
|  | 244 | NE | 0 | 0 | 0/2 |
|  | 245 | NE | 1 | 0 | 1/2 |
|  | 246 | NE | 1 | 0 | 1/2 |
|  | 247 | NE | 1 | 0 | 1/2 |
|  | 248 | NE | 0 | 0 | 0/2 |
|  | 249 | NE | 1 | 1 | 2/2 |
|  | 250 | NE | 0 | 0 | 0/2 |
|  | 251 | NE | 0 | 0 | 0/2 |
|  | 252 | NE | 0 | 0 | 0/2 |
|  | 253 | NE | 1 | 1 | 2/2 |
|  | 254 | NE | 1 | 1 | 2/2 |
|  | 255 | NE | 1 | 0 | 1/2 |
|  | 256 | NE | 0 | 0 | 0/2 |
|  | 257 | NE | NE | 1 | 1/1 |
|  | 258 | NE | 1 | 1 | 2/2 |
|  | 259 | NE | 1 | 0 | 1/2 |
|  | 260 | NE | 0 | 0 | 0/2 |
|  | 261 | NE | 0 | 0 | 0/2 |
|  | 262 | NE | 0 | 1 | 1/2 |
|  | 263 | NE | 1 | 0 | 1/2 |
|  | 264 | NE | 0 | 0 | 0/2 |
|  | 265 | NE | 0 | 0 | 0/2 |
|  | 266 | NE | 0 | 0 | 0/2 |
|  | 267 | NE | 0 | 0 | 0/2 |
| Supraclavicular LN | 268 | NE | 0 | 0 | 0/2 |
|  | 269 | NE | 1 | 0 | 1/2 |
|  | 270 | NE | 1 | 1 | 2/2 |
|  | 271 | NE | 1 | 0 | 1/2 |
|  | 272 | NE | 1 | 0 | 1/2 |
|  | 273 | NE | 0 | 0 | 0/2 |
|  | 274 | NE | NE | 1 | 1/1 |
|  | 275 | NE | 0 | 0 | 0/2 |
|  | 276 | NE | 1 | 0 | 1/2 |
|  | 277 | NE | 0 | 1 | 1/2 |
|  | 278 | NE | 1 | 1 | 2/2 |
|  | 279 | NE | 0 | 0 | 0/2 |
|  | 280 | NE | 0 | 0 | 0/2 |
|  | 281 | NE | 0 | 0 | 0/2 |
|  | 282 | NE | 0 | 0 | 0/2 |
|  | 283 | NE | 0 | 0 | 0/2 |
|  | 284 | NE | NE | 0 | 0/1 |
|  | 285 | NE | 0 | 0 | 0/2 |
|  | 286 | NE | 1 | 1 | 2/2 |
|  | 287 | NE | 0 | 0 | 0/2 |

**1 = Correct diagnosis; 0 = Incorrect diagnosis; NE = Not Examined; Yellow lines = Perfect agreement.**
| Metastatic sites | Sr. No. | 1st examiner | 2nd examiner | 3rd examiner | Score |
| --- | --- | --- | --- | --- | --- |
|  | 288 | NE | 0 | 1 | 1/2 |
|  | 289 | NE | 0 | 0 | 0/2 |
|  | 290 | NE | 1 | 1 | 2/2 |
|  | 291 | NE | 1 | 1 | 2/2 |
|  | 292 | NE | NE | 1 | 1/1 |
|  | 293 | NE | NE | 0 | 0/1 |
|  | 294 | NE | 1 | 1 | 2/2 |
|  | 295 | NE | 1 | 0 | 1/2 |
|  | 296 | NE | 1 | 1 | 2/2 |
|  | 297 | NE | 1 | 1 | 2/2 |
|  | 298 | NE | 1 | 1 | 2/2 |
| **1 = Correct diagnosis; 0 = Incorrect diagnosis; NE = Not Examined; Yellow lines = Perfect agreement.** |  |  |  |  |  |

### Ethics approval

This study was approved by the Institutional Ethics Committee of Tata Memorial Centre, Advanced Centre for Treatment, Research and Education in Cancer (Approval no. 900955).

## Results

The accuracy of prediction of the primary site of origin was 46%, 46% and 45% for the first, second and third examiner respectively. However, there was very little inter-observer agreement, and Kappa statistic generated the value of 0.27 which is much below the figure of 0.4 required to detect even a moderate level of concordance (highlighted in yellow, Table 2). In only 56 / 298 (18.8%) cases did the examiners agree with each other’s diagnosis. Thus the possibility cannot be excluded that the correct diagnoses made by the examiners had happened by play of chance.

## Discussion

To the best of our knowledge, this is the first study to blindly test the entrenched belief that a metastatic tumour is histologically similar to the primary. The results reported here raise a question mark on the veracity of such a belief and suggest that further research on the topic is called for. Perhaps another way to do this study would have been to compare paired samples of primary and metastatic tumours form the same patients. But, such a study would have been confounded by bias and would not have constituted a scientific experiment.

Metastasis is the leading cause of death from cancer, yet it is arguably the most poorly understood aspect of cancer research. In spite of decades of intensive investigation and millions of dollars spent on research, the mechanistic basis of the various steps involved in the metastatic cascade remains poorly understood^1,2^. We believe that the obstacle to progress in the field is the dogma that metastatic tumours are histologically similar to the primary, especially since the entire body of current metastasis research is founded upon such a premise. By raising an element of doubt, our study may help to energize new ideas and new avenues of research leading to better understanding of the metastatic process and development of new cancer therapies.

## Data Availability

All data produced in the present work are contained in the manuscript

## Acknowledgements

We acknowledge the help provided by the technical staff of the Pathology Department of TMH for retrieving the tissue blocks, cutting histological sections and staining the slides. We thank Mr. Ashish Pawar for his help in preparing this manuscript.

## Ethics approval statement

This study was approved by the Institutional Ethics Committee of Tata Memorial Centre, Advanced Centre for Treatment, Research and Education in Cancer which waived the requirement of informed consent (Approval no. 900955).

## Author contributions

**AS** planned and conducted the study, acquired, analyzed and reported the data; **NJ, TS** and **MR** acquired and analyzed the data; **RD** analyzed and interpreted the data and wrote the paper; **IM** conceptualized and designed the study, analyzed and interpreted the data and wrote the manuscript.

